# Knowledge, Attitude, and Practices Regarding Dengue Infection: A Community-Based Study in Rural Cox’s Bazar

**DOI:** 10.1101/2024.10.27.24316216

**Authors:** Ely Prue, Al Asmaul Husna, Shazia Rokony, Aung Sain Thowai, Shahra Tanjim Moulee, Afsana Jahan, Aysha Khatun, Moloy Sarkar, Saima Bibi, Tahsin Tasneem Tabassum, Mohammad Nurunnabi

## Abstract

**Introduction:** Dengue, an emerging infectious disease spread by mosquitoes, poses a significant public health challenge, especially in tropical and subtropical areas like Bangladesh.

**Methods:** We conducted a community-based cross-sectional study to assess the levels of knowledge, attitude, and practice (KAP) towards dengue infection among 484 adult rural people in the purposively selected Shikderpara and Muhuripara villages, Cox’s Bazar. Face-to-face interviews were performed at the convenience of study participants using a pretested, semi-structured questionnaire.

**Results:** The mean age of participants were 33.3±13.5 years, with the majority (53.9%) extending between 18 and 34 years. About two-thirds (72.9%) of the residents were literate. Most participants demonstrated average knowledge (84.3%), a positive attitude towards dengue infection (63.0%), and average prevention practices (57.2%). There was a significant association between participants’ practice levels and their knowledge and attitude (p<0.05), along with a statistically significant correlation between their knowledge and dengue prevention practices (p<0.05).

**Conclusion:** Although the attitude toward dengue infection was positive, knowledge and practices were average. This can be improved by promoting community participation and implementing comprehensive public health measures at all levels.

**Categories:** Communicable Disease, Emerging Public Health Issue.

## Introduction

Dengue fever poses a serious threat to public health, especially in tropical and subtropical areas where the main dengue virus vector, the Aedes aegypti mosquito, is abundant [1, 2]. Bangladesh is among the countries most severely affected by dengue fever, largely due to a variety of factors including its tropical climate, dense population, unplanned urbanization, inadequate vector control, and low literacy rates, among others [3,4]. A total of 5551 cases and 93 reported deaths were reported during the first official dengue outbreak in 2000 [5]. The estimated number of reported dengue cases was 2,430 in 2001, 6,232 in 2002, 3,934 in 2004, 3,162 in 2015, 6,060 in 2016, and 10,148 in 2018 [6]. During the 2020 pandemic, the Directorate General of Health in Bangladesh reported a total of 1,026 confirmed dengue cases, along with 3 confirmed dengue-related deaths [7]. In 2021, a total of 28,429 dengue cases and 105 related deaths were recorded [8]. Bangladesh experienced its major dengue outbreak in 2019, with over 100,000 reported cases and 120 deaths. This was followed by the second-largest outbreak in 2022, with 60,078 dengue cases and 266 dengue-related deaths reported as of December 10, 2022 [9,10].

Dengue is caused by the positive-sense, single-stranded RNA virus known as DENV. Four serotypes: DENNV-1, DENV-2, DENV-3, and DENV-4 belong to the Flavivirus genus within the Flaviviridae family [11,12]. Through the bites of infected female Aedes mosquitoes, mainly Aedes aegypti and, to a lesser extent, Aedes albopictus, DENV is spread through a human-mosquito-human cycle. When female mosquitoes feed on the viremic blood of a human with DENV infection, the virus spreads [13].

Humans can develop a range of illnesses from DENV, ranging from asymptomatic or mild fever to more severe conditions such as dengue hemorrhagic fever and dengue shock syndrome, which can be fatal if left untreated [8]. Patients with classical dengue typically present with fever, myalgia, arthralgia, retro-orbital pain, and rash initially. Hemorrhagic symptoms, such as sub-conjunctival hemorrhage, petechiae, and epistaxis, may also occur, either with or without accompanying shock [14,15]. Additional clinical features include respiratory symptoms, gastrointestinal disturbances, low platelet count, and abnormal liver function tests [16]. Severe plasma leakage can lead to dengue shock syndrome, fluid accumulation with respiratory distress, severe bleeding, and major organ involvement-such as liver damage (AST or ALT ≥1000), central nervous system involvement causing impaired consciousness, and complications in other organs like the heart. These severe transitions may occasionally result in death [17,18].

Despite ongoing advancements in dengue research globally, ensuring effective treatment and preventive measures remains challenging. Therefore, further research is essential to identify effective preventive strategies [19]. Although several studies have investigated public awareness, attitudes, and practices related to dengue in Bangladesh [20,21]. Therefore, the aim of this study was to assess the knowledge, attitudes, and preventive measures regarding dengue among rural residents of Cox’s Bazar, Bangladesh. The successful implementation of prevention programs largely depends on the awareness of the broader population.

## Methods

### Study design and settings

This community-based cross-sectional study was conducted to assess the knowledge, attitude, and practices regarding dengue infection (DI) among rural residents. The study sites were purposefully selected, consisting of two villages: Shikderpara and Muhuripara, located in Cox’s Bazar, Bangladesh.

### Sample selection criteria

The study included 484 adults aged 18 years and older who were available at the time of data collection. Aged ≥65 years and who had suffered from serious illness were excluded from the study.

### Data collection procedures

From July to October 2023, a pretested, face-to-face semi-structured questionnaire was utilized to interview study participants at their convenience. As part of the Residential Field Site Training (RFST), third-year medical students from Cox’s Bazar Medical College collected the data. The questionnaire included sections on the participants’ socio-demographic profiles, their knowledge about dengue infection, their attitudes toward it, and their preventive practices.

### Statistical analysis plan

Data was entered, curated, and analyzed using IBM SPSS Version 23 (New York, USA). Descriptive statistics were expressed as frequency (percentage) and mean (±standard deviation, or SD) for categorical and continuous data, respectively. The chi-square test and Fisher exact test were used to assess the significance of associations between two nominal variables. A p-value of <0.05 at a 95% confidence interval (CI) was considered significant for all statistical tests.

- **Levels of knowledge:** The participant’s knowledge of DI was scored based on 26 questions. Here, the score for an incorrect answer was ‘0’ and the score for a correct answer was ‘1’. The total range of scores was 0-26. Scores 0-9 were considered poor (<40%), 10-20 were considered average (40-70%), and 21-26 were considered good (≥80%).
- **Levels of attitude:** The participant’s attitude towards DI was scored based on 9 questions. Here, the score for an incorrect answer was ‘0’ and the score for a correct answer was ‘1’. The total range of scores was 0-9. Scores 0-3 were considered poor (<40%), 4-6 were considered average (40-70%), and 7-9 were considered good (≥80%).
- **Levels of practice:** The participant’s practice towards DI was scored based on 10 questions. Here, the score for an incorrect answer was ‘0’ and the score for a correct answer was ‘1’. The total range of scores was 0-10. Scores 0-3 were considered poor (<40%), 4-7 were considered average (40-70%), and 8-10 were considered good (≥80%).

### Ethical consideration

Participation in the study was voluntary, and confidentiality was ensured by assigning each participant an individual code number. The interviewer obtained informed written consent and permission before conducting the interviews. All procedures adhered to the guidelines of the Declaration of Helsinki. The study was approved by Cox’s Bazar Medical College, Cox’s Bazar 4700, Bangladesh. (Reference: 59.14.2200.041.19.000.23/1671)

## Results

The mean age was 33.3±13.5 year, with the majority (53.9%) ranging to the 18–34 age groups. The predominant characteristics were female (64.3%), Muslim (84.1%), and married (82.0%). In terms of education, over half of the participants (55.0%) had not completed their undergraduate studies, while a notable portion was illiterate (27.1%). Most participants identified as homemakers (56.0%) and resided in semi-building houses (55.2%). The majority hailed from nuclear families (65.1%) and earned more than 10,000 taka per month (56.4%). (Table 1)

**Table 1:**
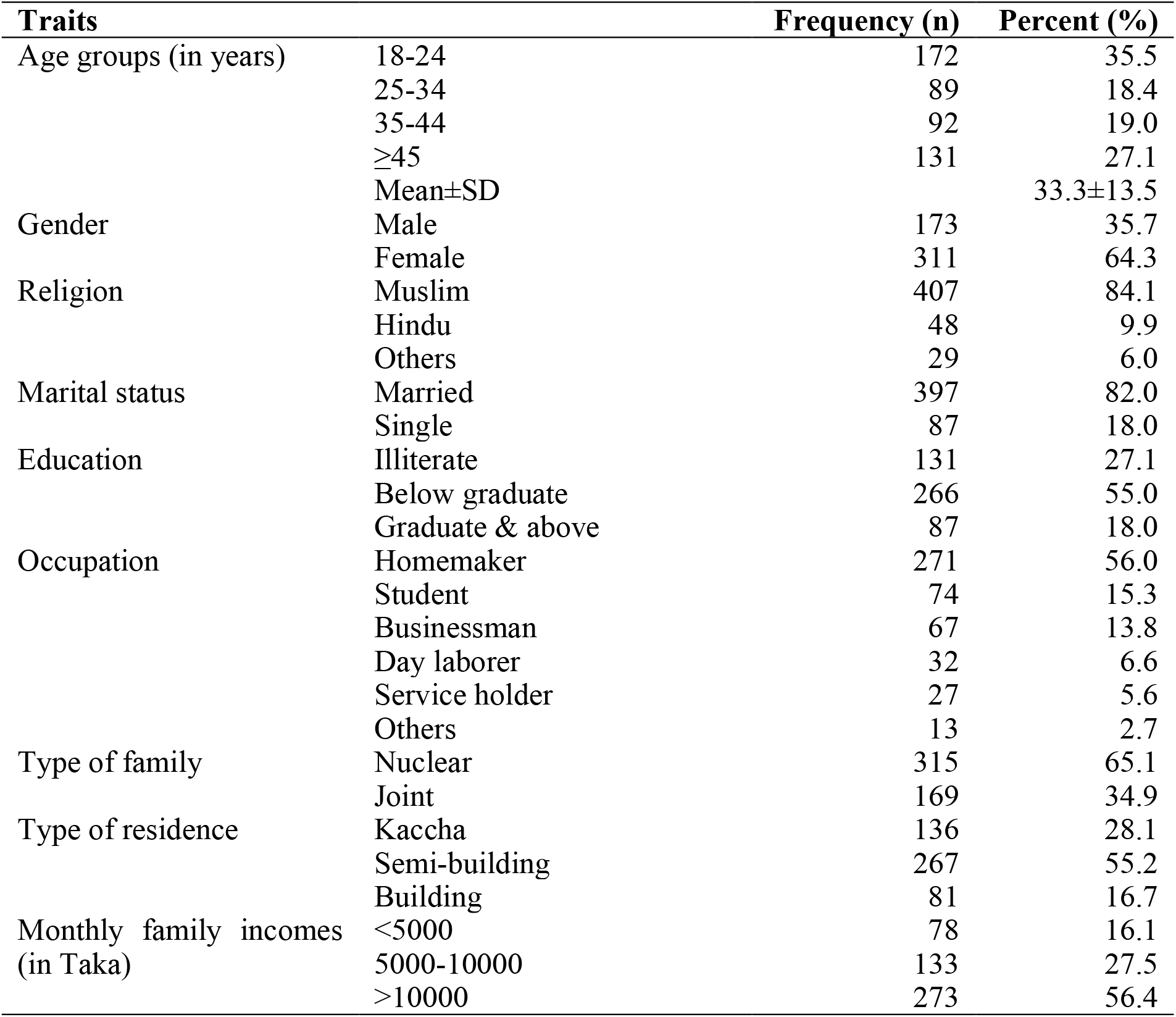
Socio-demographic characteristics of the respondents (n=484)

| **Traits** |  | **Frequency (n)** | **Percent (%)** |
| --- | --- | --- | --- |
| Age groups (in years) | 18-24 | 172 | 35.5 |
|  | 25-34 | 89 | 18.4 |
|  | 35-44 | 92 | 19.0 |
|  | ≥45 | 131 | 27.1 |
|  | Mean±SD |  | 33.3±13.5 |
| Gender | Male | 173 | 35.7 |
|  | Female | 311 | 64.3 |
| Religion | Muslim | 407 | 84.1 |
|  | Hindu | 48 | 9.9 |
|  | Others | 29 | 6.0 |
| Marital status | Married | 397 | 82.0 |
|  | Single | 87 | 18.0 |
| Education | Illiterate | 131 | 27.1 |
|  | Below graduate | 266 | 55.0 |
|  | Graduate & above | 87 | 18.0 |
| Occupation | Homemaker | 271 | 56.0 |
|  | Student | 74 | 15.3 |
|  | Businessman | 67 | 13.8 |
|  | Day laborer | 32 | 6.6 |
|  | Service holder | 27 | 5.6 |
|  | Others | 13 | 2.7 |
| Type of family | Nuclear | 315 | 65.1 |
|  | Joint | 169 | 34.9 |
| Type of residence | Kaccha | 136 | 28.1 |
|  | Semi-building | 267 | 55.2 |
|  | Building | 81 | 16.7 |
| Monthly family incomes<br>(in Taka) | <5000 | 78 | 16.1 |
|  | 5000-10000 | 133 | 27.5 |
|  | >10000 | 273 | 56.4 |

Table 2 indicates that a significant proportion of participants were aware of various aspects related to dengue infection. Specifically, 81.6% knew about dengue infection itself, while 81.4% recognized mosquitoes as the cause. Additionally, 41.7% correctly identified Aedes mosquitoes as the specific vector for dengue transmission. Most participants were knowledgeable about stagnant water being a breeding ground for mosquitoes (64.3%) and their feeding times (54.1%). A considerable number of participants acknowledged that dengue fever affects all age groups (77.9%), presents flu-like symptoms (43.8%), transmitted by direct contact (44.2%), differs from malaria (61.0%), and can be fatal (90.5%). Common preventive measures cited by participants included the use of mosquito coils/electric bats (66.5%), bed nets, and window screens (54.3%), insecticide spraying (43.0%), and eliminating stagnant water sources (42.6%). The most frequently reported symptoms of dengue infection were fever (79.1%), fatigue (75.0%), and nausea and vomiting (71.9%).

**Table 2:** Knowledge, attitude, and practices on dengue infection (n=484)

| Attributes | Positive responses |  |
| --- | --- | --- |
|  | Frequency (n) | Percent (%) |
| **Knowledge about dengue infection** |  |  |
| Knows about dengue infection | 395 | 81.6 |
| Mosquito causes dengue infection | 394 | 81.4 |
| Aedes as the mosquito causes dengue | 202 | 41.7 |
| Breeding place of mosquito (Stagnant water) | 311 | 64.3 |
| Feeding time of mosquito (Morning & afternoon) | 222 | 45.9 |
| Dengue fever affects all age | 377 | 77.9 |
| Dengue sometimes show flu like illness | 212 | 43.8 |
| Dengue fever transmits by direct contact | 214 | 44.2 |
| Dengue is different from malaria | 295 | 61.0 |
| Dengue infection causes death | 438 | 90.5 |
| Preventive measures (Mosquito coil) | 322 | 66.5 |
| Preventive measures (Using bed nets and window screens) | 263 | 54.3 |
| Preventive measures (Spraying insecticides) | 208 | 43.0 |
| Preventive measures (Eliminating stagnant water sources) | 206 | 42.6 |
| Preventive measures (Proper garbage dumping) | 188 | 38.8 |
| Preventive measures (Using mosquito repellants) | 157 | 32.4 |
| **Attitude towards dengue infection** |  |  |
| Fear of contracting dengue | 399 | 82.4 |
| Dengue infection is serious illness | 444 | 91.7 |
| Dengue infection is preventable | 432 | 89.3 |
| The government should be responsible for controlling dengue | 371 | 76.7 |
| Individually contribute to prevent dengue | 398 | 82.2 |
| Dengue requires immediate treatment because there is no cure | 357 | 73.8 |
| The public can play a crucial role in dengue control | 434 | 89.7 |
| Elimination of larvae at the breeding site is absolutely required | 438 | 90.5 |
| Dengue has a high potential to spread in the future if an outbreak occurs | 339 | 70.0 |
| **Practices towards prevent dengue infection** |  |  |
| Pattern of practices for sweeping yard (Once daily) | 401 | 82.9 |
| Pattern of practices for sweeping yard (Once in alternative day) | 199 | 41.1 |
| Measures taken to prevent DI (Used of mosquito nets) | 411 | 84.9 |
| Measures taken to prevent DI (Cleaned garbage) | 405 | 83.7 |
| Measures taken to prevent DI (Covered water container at home) | 404 | 83.5 |
| Measures taken to prevent DI (Utilized mosquito repellent products) | 401 | 82.9 |
| Measures taken to prevent DI (Disposed of water-holding containers) | 377 | 77.9 |
| Measures taken to prevent DI (Covered the body with clothes) | 319 | 65.9 |
| Measures taken to prevent DI (Used smoke to repel mosquitoes) | 261 | 53.9 |
| Measures taken to prevent DI (Used window screens) | 253 | 52.3 |

The majority of participants held a positive attitude towards dengue infection. Specifically, 82.4% expressed fear of contracting dengue. Moreover, most participants perceived dengue as a serious illness (91.7%), preventable (89.3%), felt the government should take responsibility for its control (76.7%). Additionally, a significant portion believed in individual contribution to dengue prevention (82.2%), recognized the necessity of immediate treatment for dengue (73.8%), acknowledged the public’s crucial role in dengue control (89.7%), emphasized the absolute necessity of eliminating larvae at breeding sites (90.5%), and 70.0% expressed concerns about the potential future spread of dengue (Table 2).

The practices adopted by participants to prevent dengue infection. The majority of the participants stated sweeping their yards daily (82.9%). Moreover, common preventive measures included the use of mosquito nets (84.9%), cleaning of garbage (83.7%), covering water containers at home (83.5%), utilizing mosquito repellent products (82.9%), disposing of water-holding containers (77.9%), covering the body with clothes (65.9%), using smoke to repel mosquitoes (53.9%), and using window screens (52.3%). (Table 2)

The participants mean knowledge score was 14.9 (SD: 4.1; range 0-26), indicating that the majority (84.3%) possessed an average level of knowledge about dengue infection. Regarding attitudes, the mean score was 6.8 (SD: 1.3; range 0-9), with a significant portion (63.0%) demonstrating a good attitude towards dengue infection. In terms of practices, the mean score was 7.1 (SD: 1.7; range 0-10), with the majority (57.2%) exhibiting average practices in preventing dengue infection. (Table 3)

**Table 3:** Levels and scores of knowledge, attitude, and practice towards dengue infection (n=484)

| Levels |  | Frequency (n) | Percent (%) |
| --- | --- | --- | --- |
| Knowledge | Poor | 39 | 8.1 |
|  | Average | 408 | 84.3 |
|  | Good | 37 | 7.6 |
|  | Mean±SD |  | 14.9±4.1 |
| Attitude | Poor | 12 | 2.5 |
|  | Average | 167 | 34.5 |
|  | Good | 305 | 63.0 |
|  | Mean±SD |  | 6.8±1.3 |
| Practices | Poor | 7 | 1.4 |
|  | Average | 277 | 57.2 |
|  | Good | 200 | 41.3 |
|  | Mean±SD |  | 7.1±1.7 |

Participants’ levels of knowledge showed significant associations with their age, gender, religion, marital status, educational level, occupation, and type of residence (p<0.05). There were a significant association between participants’ attitude levels and their age, gender, religion, marital status, educational level, occupation, type of family, type of residence, family income, and their levels of knowledge (p<0.05). Furthermore, there were also a significant association between participants’ practice levels and their age, religion, marital status, educational level, occupation, type of residence, and family income, along with their levels of knowledge and attitude (p<0.05). (Table 4 and 5)

**Table 4:** Association of the levels of knowledge, attitude, and practice with socio-demographic characteristics (n=484)

| Variables | Levels of knowledge |  |  | p-value | Levels of attitude |  |  | p-value | Levels of practice |  |  | p-value |
| --- | --- | --- | --- | --- | --- | --- | --- | --- | --- | --- | --- | --- |
|  | Poor | Average | Good |  | Poor | Average | Good |  | Poor | Average | Good |  |
|  | n(%) | n(%) | n(%) |  | n(%) | n(%) | n(%) |  | n(%) | n(%) | n(%) |  |
| Age groups (in years) |  |  |  |  |  |  |  |  |  |  |  |  |
| 15-24 | 5(2.9) | 153(89.0) | 14(8.1) | 0.001 | 4(2.3) | 78(45.3) | 90(53.3) | 0.000 | 3(1.7) | 78(45.3) | 91(52.9) | 0.000 |
| 25-34 | 2(2.2) | 80(89.9) | 7(7.9) |  | 3(3.4) | 14(15.7) | 72(80.9) |  | 1(1.1) | 49(55.1) | 39(43.8) |  |
| 35-44 | 15(16.3) | 67(72.8) | 10(10.9) |  | 1(1.1) | 25(27.2) | 66(71.7) |  | 0(0.0) | 76(82.6) | 16(17.4) |  |
| ≥45 | 17(13.0) | 108(82.4) | 6(4.6) |  | 4(3.1) | 50(38.2) | 77(58.8) |  | 3(2.3) | 74(56.5) | 54(41.2) |  |
| Gender |  |  |  |  |  |  |  |  |  |  |  |  |
| Male | 12(6.9) | 139(80.3) | 22(12.7) | 0.007 | 11(6.4) | 69(39.9) | 93(53.8) | 0.000 | 5(2.9) | 99(57.2) | 69(39.9) | 0.168 |
| Female | 27(8.7) | 269(86.5) | 15(4.8) |  | 1(0.3) | 98(31.5) | 212(68.2) |  | 2(0.6) | 178(57.2) | 131(42.1) |  |
| Religion |  |  |  |  |  |  |  |  |  |  |  |  |
| Muslim | 27(6.6) | 347(85.3) | 33(8.1) | 0.030 | 9(2.2) | 150(36.9) | 248(60.9) | 0.041 | 6(1.5) | 218(53.6) | 183(45.0) | 0.012 |
| Hindu | 5(10.4) | 41(85.4) | 2(4.2) |  | 3(6.3) | 12(25.0) | 33(68.8) |  | 0(0.0) | 36(75.0) | 12(25.0) |  |
| Others | 7(24.1) | 20(69.0) | 2(6.9) |  | 0(0.0) | 5(17.2) | 24(82.8) |  | 1(3.4) | 23(79.3) | 5(17.2) |  |
| Marital status |  |  |  |  |  |  |  |  |  |  |  |  |
| Married | 34(8.6) | 339(85.4) | 24(6.0) | 0.015 | 8(2.0) | 149(37.5) | 240(60.5) | 0.007 | 4(1.0) | 216(54.4) | 177(44.6) | 0.004 |
| Single | 5(5.7) | 69(79.3) | 13(14.9) |  | 4(4.6) | 18(20.7) | 65(74.7) |  | 3(3.4) | 61(70.1) | 23(26.4) |  |
| Education |  |  |  |  |  |  |  |  |  |  |  |  |
| Illiterate | 18(13.7) | 107(81.7) | 6(4.6) | 0.004 | 0(0.0) | 52(39.7) | 79(60.3) | 0.000 | 1(0.8) | 79(60.3) | 51(38.9) | 0.020 |
| Below graduate | 20(7.5) | 226(85.0) | 20(7.5) |  | 12(4.5) | 75(28.2) | 179(67.3) |  | 3(1.1) | 161(60.5) | 102(38.3) |  |
| Graduate & above | 1(1.1) | 75(86.2) | 11(12.6) |  | 0(0.0) | 40(46.0) | 47(54.0) |  | 3(3.4) | 37(42.5) | 47(54.0) |  |
| Occupation |  |  |  |  |  |  |  |  |  |  |  |  |
| Homemaker | 25(9.2) | 234(86.3) | 12(4.4) | 0.000 | 2(0.7) | 94(34.7) | 175(64.6) | 0.000 | 2(0.7) | 148(54.6) | 121(44.6) | 0.000 |
| Student | 3(4.1) | 62(83.8) | 9(12.2) |  | 5(6.8) | 9(12.2) | 60(81.1) |  | 0(0.0) | 56(75.7) | 18(24.3) |  |
| Businessman | 0(0.0) | 53(79.1) | 14(20.9) |  | 2(3.0) | 43(64.2) | 22(32.8) |  | 0(0.0) | 26(38.8) | 41(61.2) |  |
| Day laborer | 6(18.8) | 26(81.3) | 0(0.0) |  | 3(9.4) | 14(43.8) | 15(46.9) |  | 0(0.0) | 24(75.0) | 8(25.0) |  |
| Service holder | 4(14.8) | 21(77.8) | 2(7.4) |  | 0(0.0) | 6(22.2) | 21(77.8) |  | 3(11.1) | 14(51.9) | 10(37.0) |  |
| Others | 1(7.7) | 12(92.3) | 0(0.0) |  | 0(0.0) | 1(7.7) | 12(92.3) |  | 2(15.4) | 9(69.2) | 2(15.4) |  |
| Type of family |  |  |  |  |  |  |  |  |  |  |  |  |
| Nuclear | 25(7.9) | 268(85.1) | 22(7.0) | 0.744 | 12(3.8) | 105(33.3) | 198(62.9) | 0.030 | 2(0.6) | 184(58.4) | 129(41.0) | 0.127 |
| Joint | 14(8.3) | 140(82.8) | 15(8.9) |  | 0(0.0) | 62(36.7) | 107(63.3) |  | 5(3.0) | 93(55.0) | 71(42.0) |  |
| Type of residence |  |  |  |  |  |  |  |  |  |  |  |  |
| Kaccha | 24(17.6) | 109(80.1) | 3(2.2) | 0.000 | 3(2.2) | 24(17.6) | 109(80.1) | 0.000 | 3(2.2) | 79(58.1) | 54(39.7) | 0.000 |
| Semi-pucca | 10(3.7) | 232(86.9) | 25(9.4) |  | 9(3.4) | 126(47.2) | 132(49.4) |  | 1(0.4) | 138(51.7) | 128(47.9) |  |
| Pucca | 5(6.2) | 67(82.7) | 9(11.1) |  | 0(0.0) | 17(21.0) | 64(79.0) |  | 3(3.7) | 60(74.1) | 18(22.2) |  |
| Family incomes (in Taka) |  |  |  |  |  |  |  |  |  |  |  |  |
| <5000 | 5(6.4) | 70(89.7) | 3(3.8) | 0.463 | 3(3.8) | 37(47.4) | 38(48.7) | 0.046 | 3(3.8) | 44(56.4) | 31(39.7) | 0.000 |
| 5000-10000 | 14(10.5) | 108(81.2) | 11(8.3) |  | 4(3.0) | 38(28.6) | 91(68.4) |  | 0(0.0) | 111(83.5) | 22(16.5) |  |
| >10000 | 20(7.3) | 230(84.2) | 23(8.4) |  | 5(1.8) | 92(33.7) | 176(64.5) |  | 4(1.5) | 122(44.7) | 147(53.8) |  |

Chi-square test and Fisher exact test done

**Table 5:**
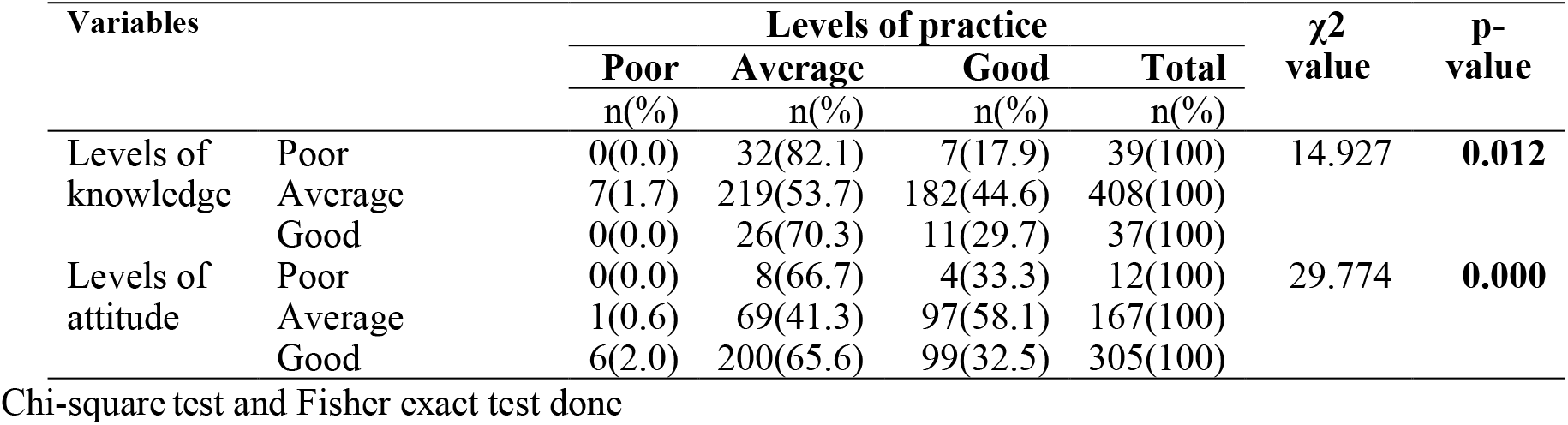
Association of the levels of knowledge, and attitude with levels of practice (n=484)

There was a significant correlation between participants’ knowledge and practices regarding DI (p<0.05). (Figure 2)

**Figure 1:**
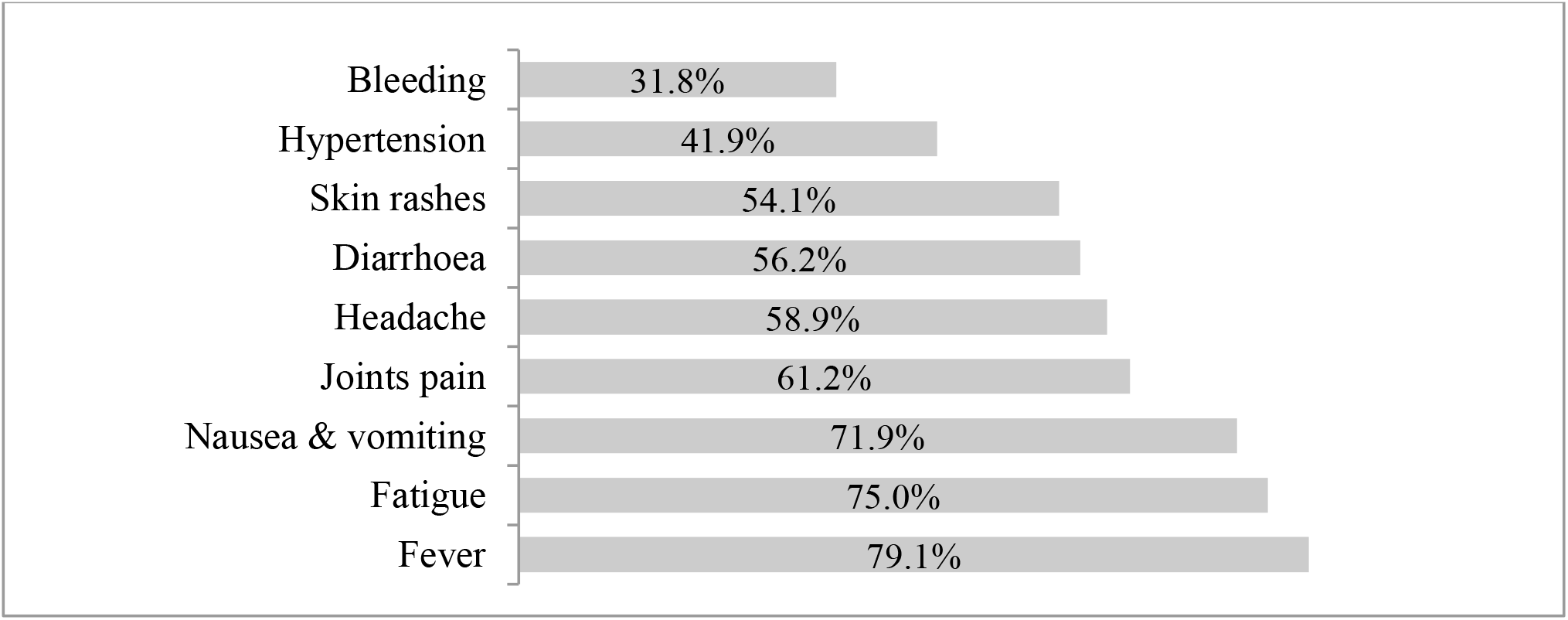
Knowledge about symptoms of dengue infection (n=484)

**Figure 2:**
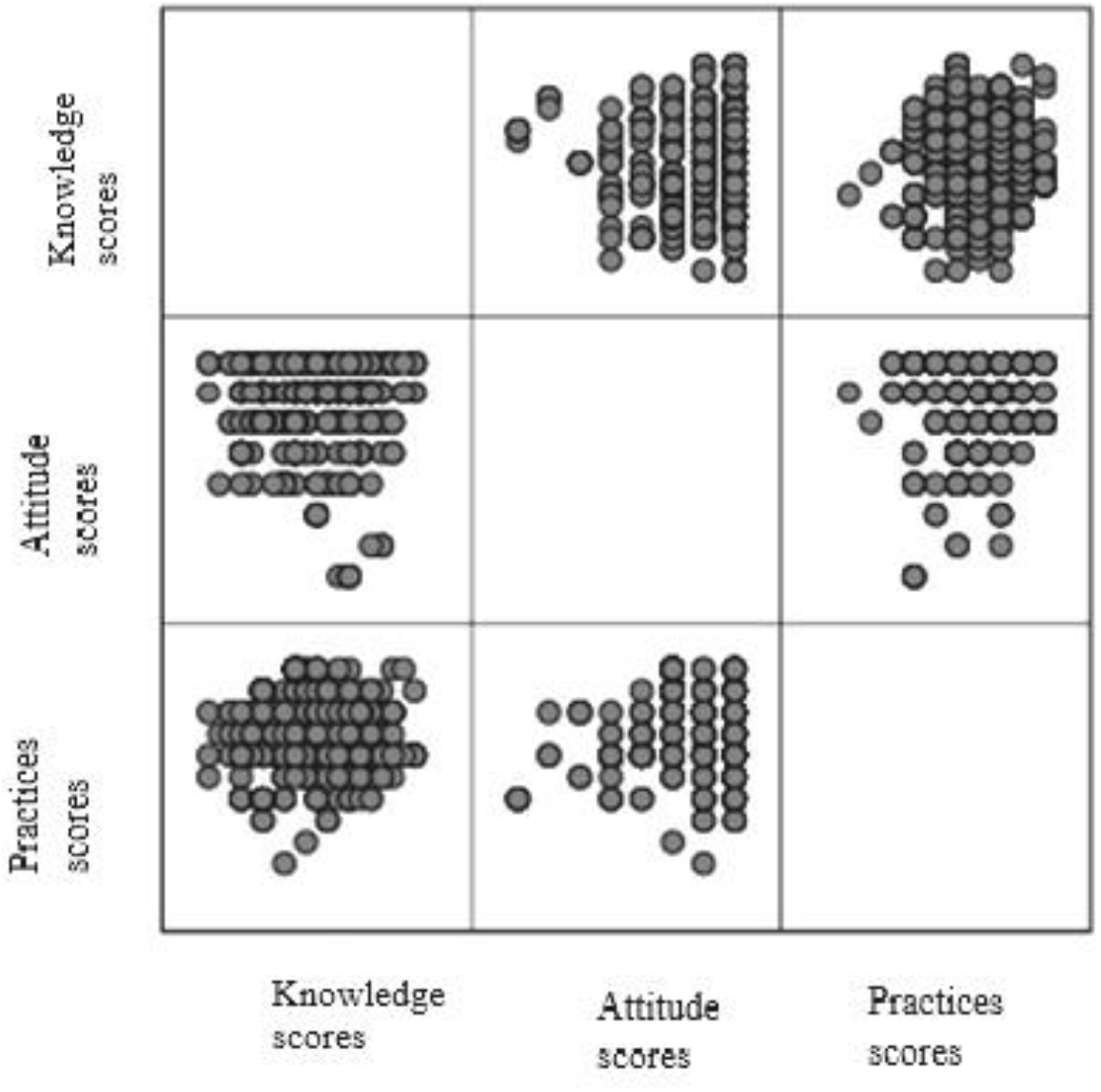
Correlations within knowledge, attitude and practice scores (n=484)

## Discussion

The majority (53.9%) of participants were in the 18–34 age groups, indicating that most were young adults. This trend was also observed in studies conducted in Bangladesh and India, where most interviewed participants were young adults [12,22]. In terms of education, a significant portion of the participants were illiterate (27.1%), slightly higher than the national illiteracy rate of 24.4% [12,23].

The majority of participants (84.3%) had an average level of knowledge of dengue infection, according to their mean knowledge score of 14.9 (SD: 4.1). Regarding attitudes, the average score was 6.8 (SD: 1.3), and 63.0% of respondents showed a positive attitude regarding dengue infection. Regarding practices, the majority (57.2%) showed an average practices in preventing dengue infection, with a mean score of 7.1 (SD: 1.7). Regarding dengue infection, it found that a moderate level of knowledge, and practices among the rural residents of Bangladesh [24]. Conversely, both urban and rural residents were found to have a good level of attitude [12,24]. This may be attributed to varying levels of literacy among the participants.

Participants’ knowledge levels were significantly associated with their age, gender, religion, marital status, educational level, occupation, and type of residence (p<0.05). Similarly, participants’ attitude levels were significantly linked to their age, gender, religion, marital status, educational level, occupation, family type, type of residence, family income, and knowledge levels (p<0.05). Additionally, significant associations were found between participants’ practice levels and their age, religion, marital status, educational level, occupation, type of residence, family income, as well as their knowledge and attitude levels (p<0.05). Furthermore, there was a statistically significant correlation between participants’ knowledge and their practices regarding DI (p<0.05). Moreover, people’s dengue prevention practices were found to be significantly influenced by their level of knowledge. This highlights the urgent need for expanded educational outreach to raise public awareness about dengue and promote preventive practices in rural communities of Bangladesh [15,24].

## Conclusion

The study revealed that the level of knowledge and practices regarding dengue infection were average or moderate, but their level of attitude was good. Efforts to enhance knowledge and practices related to dengue infection through education and comprehensive public health initiatives by local administrations are crucial for achieving more sustainable outcomes.

## Data Availability

All data produced in the present work are contained in the manuscript

## Author’s contribution

Conceptualization, methods and literature reviews: Prue E, Tabassum TT, Bibi S and Nurunnabi M; Data collection: Prue E, Husna AA; Statistical analysis: Nurunnabi M; Draft manuscript: Prue E, Husna AA, Rokony S, Thowai AS, Moulee ST, Jahan A, Khatun A, Sarkar M, Bibi S, Tabassum TT, and Nurunnabi M. All the authors worked and approved the final manuscript.

## Acknowledgments

We would like to express our gratitude to the local authorities for granting us permission to conduct our research in the communities. We also thank the participants and students for their valuable contributions to the study.

